# Estimating The Effect Of Adhering To Canada’s Food Guide 2019 Recommendations On Health Outcomes In Older Adults: A Target Trial Emulation Protocol

**DOI:** 10.1101/2024.05.29.24308054

**Authors:** Didier Brassard, Nancy Presse, Stéphanie Chevalier

## Abstract

**Background:** The Canada’s Food Guide 2019 (CFG) provides universal recommendations to individuals aged 2 years or older. The extent to which these recommendations are appropriate for older adults is unknown. Although ideal, conducting a large randomized controlled trial is unrealistic in the short term. An alternative is the target trial emulation framework for causal inference, a novel approach to improve the analysis of observational data.

**Objectives:** Our aim is to describe the protocol of a target trial emulation in older adults with emphasis on key aspects of a hypothetical sustained diet and physical activity intervention.

**Methods:** To emulate the target trial, non-experimental data from the NuAge longitudinal study (n=1753, adults aged 67 years or older) will be used. NuAge includes 4 yearly measurements of dietary intakes, covariates and outcomes. The per protocol causal contrast will be the primary causal contrast of interest to account for non-adherence. The sustained intervention strategy will be modelled using the parametric g-formula. In the hypothetical trial, participants would be instructed to meet sex-specific minimal intakes for vegetables and fruits, whole grains, animal- and plant-based protein foods, milk & plant-based beverages and unsaturated fats. Eligibility criteria, follow-up, intervention, outcomes, and causal contrast will be similar in the emulation to the target trial except for minor modifications. We will attempt to emulate randomization of treatment by adjusting for baseline covariates and pre-baseline dietary habits.

**Results:** Data collection for the NuAge study was completed in June 2008. For the present work, the main analysis has started in May 2024. Submission of the manuscript is expected by December 2024.

**Conclusion:** Emulating a target trial will provide the first evidence of the adequacy of CFG 2019 recommendations for older adults in relation to health outcomes.

## Introduction

The latest edition of Canada’s Food Guide (CFG) was published in 2019 [1]. Compared with the previous edition, key changes include the removal of the pre-specified number of servings to consume each day, a shift towards qualitative (“eat plenty of…”) instead of quantitative recommendations and the provision of universal recommendations instead of age and sex-specific recommendations. In addition, CFG recommendations primarily aim at reducing chronic disease risk. Indeed, the evidence behind CFG’s recommendations concern mitigation of cardiovascular disease, cancer and type 2 diabetes risk [2]. However, evidence from a nationally representative survey of adults aged 65 years or older from Canada suggested that greater adherence to recommendations was insufficient to meet calcium, vitamin D and folate requirements [3]. In Canada, one third of community-dwelling older adults are at high nutrition risk [4,5], highlighting the importance of maintaining adequate nutritional status in this stratum of the population. Similarly, in absence of specific recommendations on the amount of protein foods to eat, older adults may be eating less protein than required to maintain muscle mass [6–8]. Canada’s Food Guide also provide brief physical activity recommendation, but without explicit acknowledgement of the importance of these recommendations for older adults [1]. Indeed, performing a minimal amount of physical activity is paramount to maintaining muscle mass [6,7]. Thus, the universal recommendations in CFG may not be appropriate for older adults since they face unique challenges to consuming a healthy diet [9] and may require specific nutritional strategies [10].

Ideally, a randomized controlled trial (RCT) would be conducted to investigate the adequacy of CFG recommendations in older adults. However, such RCT is unlikely to be conducted in the short term. An alternative is the target trial emulation framework for causal effect estimation using observational data [11–14]. Informally, the target trial emulation framework is intended to highlight and address design issues of observational data analysis that aims to estimate a causal effect by explicitly emulating a hypothetical trial [11]. In nutritional epidemiology, common issues with design and analyses can yield results that are largely inconsistent with those of randomized trials [15]. For example, the lack of consideration of the compositional nature of diet can dramatically influence effect estimates, i.e., that increasing the intake of one food must compensated by decreasing the intake of another food in substitution modeling [15–17]. Furthermore, diet is a lifelong sustained exposure. In an observational study, the effect of diet assessed at a given time may actually reflect the cumulative exposure to prior dietary habits. In turn, ignoring prior dietary habits may result in a misalignment of “time zero” [18], since dietary habits are not randomly assigned at the beginning of the observational study. In other words, ignoring prior dietary habits makes it impossible to distinguish the effect due to prospective (hypothetical) dietary modification from the effect due to retrospective dietary habits. The target trial framework is a helpful tool to highlight and address common issues in nutritional epidemiology. Ultimately, a successful emulation of the target trial based on observational data could yield effect estimates that more closely align with those of a hypothetical future RCT. Example applications of the target trial framework include the emulation of interventions on diet [19–22], physical activity [23], or both [24].

To the best of our knowledge, the target trial framework has not been used to assess the effect of adhering to CFG’s recommendations. Accordingly, the objectives of the present study are, first, to describe the protocol of the emulation of a target trial using observational data from the NuAge study [25], a cohort of 1753 adults aged 67 to 84 years at baseline, and, second, to address key aspects of the target trial emulation in the context of a sustained lifestyle intervention strategy involving diet and physical activity. Key aspects are the description of the sustained lifestyle intervention strategy, the attempt to emulate randomization, as well as assumptions and limitations specific to diet intervention. Of note, more general introductory texts to the target trial framework are available elsewhere [11,12,14].

## Methods

### Research Question and Hypothesis

Explicitly acknowledging the causal nature of a research question is prerequisite to causal effect estimation using observational data [26–28]. The general objective of this study will be to examine the adequacy of CFG’s universal dietary recommendations for older adults. Expressed as a counterfactual statement, our objective is to answer the following question:

> *What would be the difference in a given health outcome at the end of the follow-up, if all eligible participants had increased their adherence to CFG recommendations on healthy food choices compared with, instead, if they had maintained their habitual diet?*

Specifically, among adults aged 67 to 84 years followed over 3 years, and compared with maintenance of habits, we aim to:

1. Estimate the causal effect of adhering to CFG’s dietary recommendations on markers of muscle health (e.g., physical function, muscle strength), general health (e.g., waist circumference, blood pressure, glucose), and cognitive health (i.e., Modified Mini-Mental State Exam).

*Hypothesis: adhering to recommendations positively influence general and cognitive health but has no influence on muscle health*.

2. Estimate the causal effect of adhering to a *reformulation* of CFG’s dietary recommendations, including more protein foods and a minimal physical activity recommendation, to amplify the positive health effects.

*Hypothesis: increasing the consumption of protein-rich foods positively influence muscle health, and meeting minimal physical activity recommendation (30 minutes or more per day) further amplifies positive health effects*.

## Study Design and Sample

Data from the NuAge prospective cohort study will be used to emulate the target trial [25]. The NuAge cohort comprised 1753 generally healthy community-dwelling adults aged 67 to 84 years at baseline and followed over 3 years. The baseline and each annual follow-up included comprehensive assessment on sociodemographic data, diet, physical activity, functional status as well as physical and mental health [25].

The NuAge study sample is relevant to our research question. The target population of CFG recommendations comprise all individuals aged 2 years or older, which is compatible with the target sample of the NuAge study of generally healthy older adults from the greater Montreal, Sherbrooke and Laval areas, the province of Quebec, Canada.

## Target trial

The target trial framework has been suggested as a potential solution to improve the analysis of nutritional epidemiology studies aiming at causal inference [13]. Informally, the target trial framework helps to align the observational data analysis with that of a hypothetical trial. This framework is appropriate for the research question in the present study, since we aim to estimate the effect of adhering to a hypothetical diet and physical activity intervention using on observational data. The first step of a target trial emulation is the description of the target trial, i.e., the protocol of the hypothetical randomized controlled trial we would like to conduct [11,14]. The second step is the emulation, i.e., describing how to target trial is emulated and conducting the study described in the present protocol.

**Table 1** presents the target trial and its emulation using observational data from the NuAge study. Key differences between the target trial and its emulation are that participants will be required to provide complete dietary assessment and covariate data at baseline (eligibility component); and that we will attempt to emulate the randomized assignment by adjusting for dietary intakes before baseline as well as baseline covariates (assignment component).

**Table 1:**
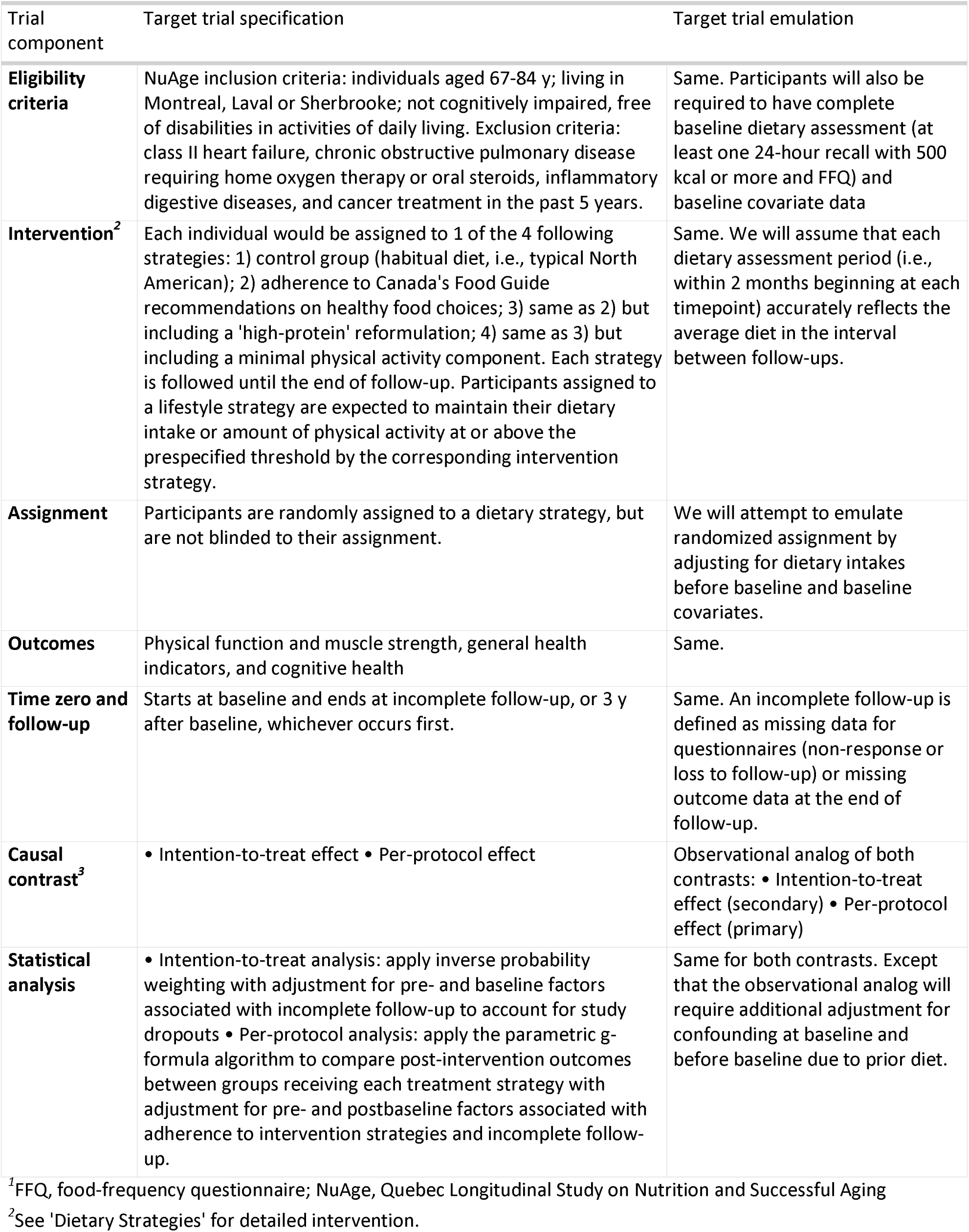
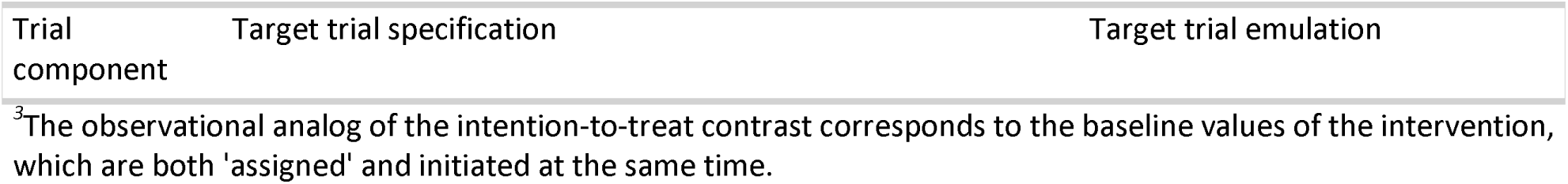
Emulation of a dietary intervention target trial using observational data from the NuAge study

| Trial component | Target trial specification | Target trial emulation |
| --- | --- | --- |
| **Eligibility criteria** | NuAge inclusion criteria: individuals aged 67-84 y; living in Montreal, Laval or Sherbrooke; not cognitively impaired, free of disabilities in activities of daily living. Exclusion criteria: class II heart failure, chronic obstructive pulmonary disease requiring home oxygen therapy or oral steroids, inflammatory digestive diseases, and cancer treatment in the past 5 years. | Same. Participants will also be required to have complete baseline dietary assessment (at least one 24-hour recall with 500 kcal or more and FFQ) and baseline covariate data |
| **Intervention<sup>2</sup>** | Each individual would be assigned to 1 of the 4 following strategies: 1) control group (habitual diet, i.e., typical North American); 2) adherence to Canada's Food Guide recommendations on healthy food choices; 3) same as 2) but including a 'high-protein' reformulation; 4) same as 3) but including a minimal physical activity component. Each strategy is followed until the end of follow-up. Participants assigned to a lifestyle strategy are expected to maintain their dietary intake or amount of physical activity at or above the prespecified threshold by the corresponding intervention strategy. | Same. We will assume that each dietary assessment period (i.e., within 2 months beginning at each timepoint) accurately reflects the average diet in the interval between follow-ups. |
| **Assignment** | Participants are randomly assigned to a dietary strategy, but are not blinded to their assignment. | We will attempt to emulate randomized assignment by adjusting for dietary intakes before baseline and baseline covariates. |
| **Outcomes** | Physical function and muscle strength, general health indicators, and cognitive health | Same. |
| **Time zero and follow-up** | Starts at baseline and ends at incomplete follow-up, or 3 y after baseline, whichever occurs first. | Same. An incomplete follow-up is defined as missing data for questionnaires (non-response or loss to follow-up) or missing outcome data at the end of follow-up. |
| **Causal contrast<sup>3</sup>** | • Intention-to-treat effect • Per-protocol effect | Observational analog of both contrasts: • Intention-to-treat effect (secondary) • Per-protocol effect (primary) |
| **Statistical analysis** | • Intention-to-treat analysis: apply inverse probability weighting with adjustment for pre- and baseline factors associated with incomplete follow-up to account for study dropouts • Per-protocol analysis: apply the parametric g-formula algorithm to compare post-intervention outcomes between groups receiving each treatment strategy with adjustment for pre- and postbaseline factors associated with adherence to intervention strategies and incomplete follow-up. | Same for both contrasts. Except that the observational analog will require additional adjustment for confounding at baseline and before baseline due to prior diet. |

<sup>1</sup>FFQ, food-frequency questionnaire; NuAge, Quebec Longitudinal Study on Nutrition and Successful Aging
<sup>2</sup>See 'Dietary Strategies' for detailed intervention.

| Trial component | Target trial specification | Target trial emulation |
| --- | --- | --- |

<sup>3</sup>The observational analog of the intention-to-treat contrast corresponds to the baseline values of the intervention, which are both 'assigned' and initiated at the same time.

Each part of the target trial and its emulation are described below.

### Eligibility Criteria

The inclusion criteria for the target trial are the same as for the NuAge study [25]. In the emulation, participants will be required to have at least one 24-hour dietary recall completed at baseline with 500 calories or more, as well as complete covariate data at baseline, as identified below.

### Hypothetical Interventions

The hypothetical intervention strategies evaluated will be:

1. no change in dietary habits or physical activity (“control” intervention);
2. adherence to CFG’s recommendations;
3. adherence to CFG’s recommendations including reformulation (i.e., higher intake of protein foods);
4. adherence to CFG’s recommendations including reformulation (i.e., higher intake of protein foods) and performing at least 30 minutes of aerobic physical activity.

Physical activity recommendations are not traditionally at the forefront of CFG’s recommendations. However, CFG does mention that “*at least 150 minutes of moderate-to vigorous-intensity aerobic physical activity per week […] is recommended to achieve health benefits*” [1]. Thus, recognizing the key role of exercise to maintaining health and muscle for older adults, the 4^th^ hypothetical intervention includes a formal physical activity recommendation. In the target trial, the physical activity corresponds to performing aerobic exercise of light to vigorous-intensity aerobic for at least 30 minutes per day [6].

#### The challenge of a well-defined nutritional intervention

Emulating a well-defined dietary intervention for CFG’s recommendations is challenging. First, recommendations in the latest edition of CFG are qualitative and flexible (e.g., “*Eat plenty of vegetables and fruits*”; [1,29]). Thus, multiple suitable, but nonetheless very different, dietary patterns can achieve adherence to recommendations. Second, CFG’s recommendations target both food intakes (e.g., vegetables and fruits) and nutrients (e.g., saturated fats). The nutrient-based recommendations can be achieved by intervening on the consumption of multiple different foods. For example, to achieve the hypothetical intervention “*decreasing consumption of calories from saturated fats*” one could decrease saturated fats from dairy, nuts and low nutritive value foods altogether. Arguably, the relationship between these food categories and a given health outcome may vary greatly.

To estimate a causal effect using observational data, the hypothetical interventions must be sufficiently well-defined or that until “no meaningful variation” of that intervention remains [30,31]. In other words, the hypothetical diet interventions should be elaborated until no additional dietary characteristics are deemed impactful in regards of the outcome of interest. Another consideration is that the modelling of hypothetical interventions should ideally be conducted with dietary intakes expressed using the same units. For example, mixing food intakes expressed in servings and grams in a statistical model may cause poor estimation of causal effects [17]. Finally, the statistical approach used to account for ‘total energy’ or total food intake also affects the causal effect of interest and should be consistent with the research question [17,31–33].

#### Diet Simulations

For the present study, adherence to CFG’s recommendations was defined based on simulated diets generated by Health Canada [34] and summarized in **Supplemental Table 1**. The simulated diets were designed to meet both CFG’s recommendations on healthy food choices and nutrient requirements (Dietary Reference Intake).

These diets achieve near perfect Healthy Eating Food Index (HEFI)-2019 scores (>78/80) through relatively high intakes of recommended foods (i.e., vegetables and fruits, whole-grain foods, protein foods, and unsweetened milk and plant-based beverages with protein) and null intakes of foods not recommended (i.e., non-whole grain foods, other low nutritive value foods, juice, sugary drinks and alcohol, and fatty foods rich in saturated fats). The HEFI-2019 score indicates the extent to which dietary intakes are consistent with CFG’s recommendations on healthy food choices [29,35].

Note that the HEFI-2019 could have been used as main exposure to measure adherence to CFG. However, the use of a composite score metric would not fully satisfy the criterion of a well-defined intervention to estimate a causal effect. First, high HEFI-2019 scores, and high adherence to CFG’s recommendations, can be achieved through many different strategies or dietary patterns. In the context of observational data, the specific strategies through which individuals achieve high HEFI-2019 score would be based on dietary habits and patterns self-selected by the participants. This approach is similar to asking hypothetical trial participants to modify their intakes without clearly indicating how, which would obscure the causal effect estimated. Second, the HEFI-2019 score includes recommendations both on foods and on nutrients. As described above, mixing servings and grams in a statistical models may cause poor estimation of causal effects [17].

We stress that the diets simulated by Health Canada were not actually consumed by older adults. For this reason, the simulated values for vegetables and fruits, whole-grain foods and plant-based protein foods *exceed* the 99^th^ percentile of the distribution of usual intakes of these food categories as estimated in adults aged 65 years or more from the Canadian Community Health Survey 2015 -Nutrition [3]. In **Table 2**, the target intakes vegetables and fruits, whole grains, and plant-based protein foods for the adhering to Canada’s Food Guide 2019 recommendations intervention were revised to correspond, at most, to the 90^th^ percentile of the distribution of usual intakes among Canadians aged 65 years or more in 2015 [3]. Since Canada’s Food Guide 2019 does not have a portion size system, reference amounts (RA) were used as a proxy for servings. RA are regulated quantity of foods that reflect the portion size typically consumed at 1 sitting in Canada. RA were used by Health Canada to simulate diet consistent with Canada’s Food Guide 2019 recommendations and Dietary Reference Intake (Supplemental Table 1), hence RAs are adequate for the present work.

**Table 2:**
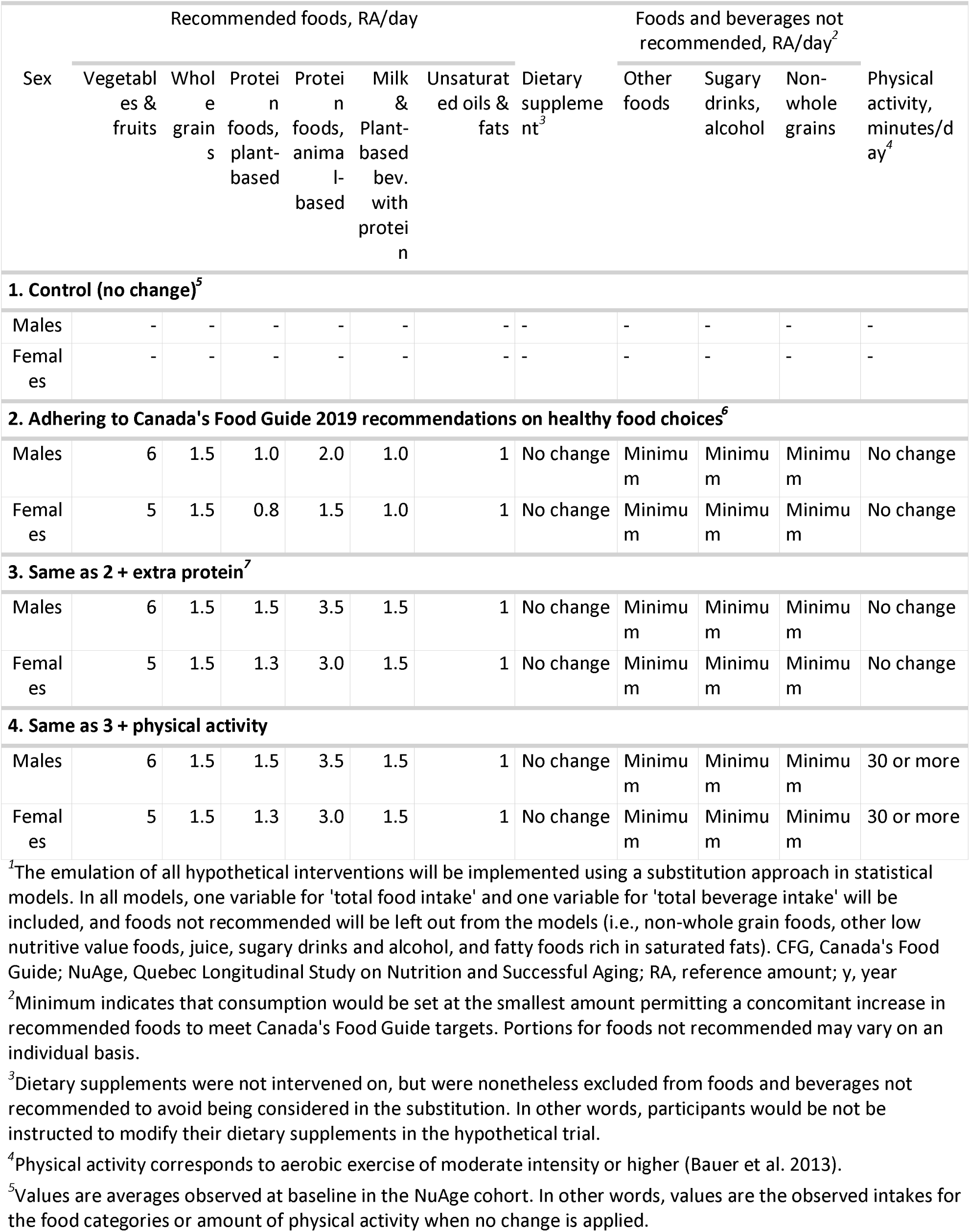
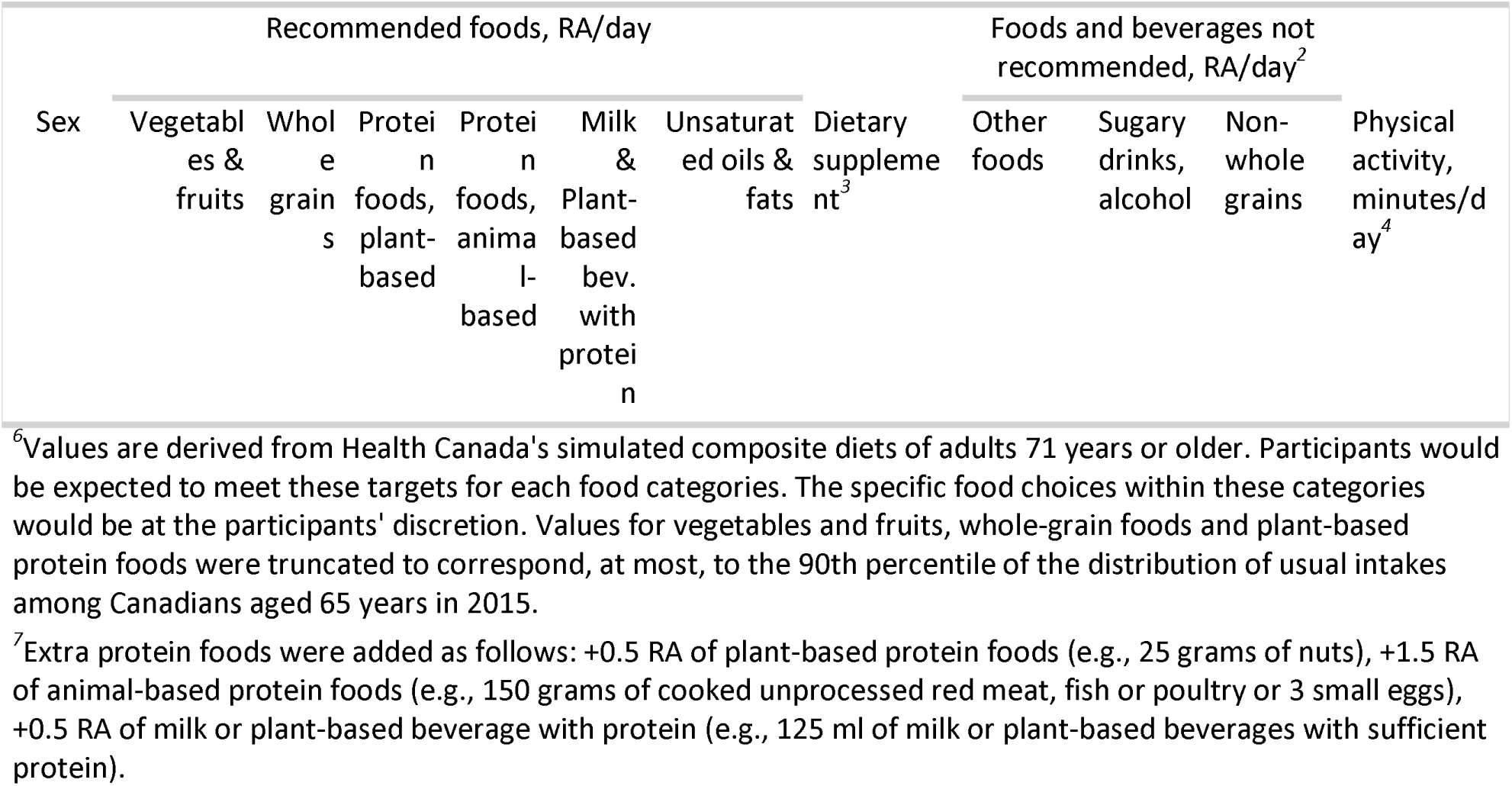
Hypothetical diet and exercise interventions emulated in the NuAge cohort study, by sex

#### Implementation

In the target trial, the sustained intervention strategy could be implemented as follows:

1. the participant’s usual dietary intakes and physical activity would be assessed by research dietitians at each study visit;
2. if reported food intakes and duration of physical activity were **equal to or above** the prespecified thresholds (Table 2), no change would be suggested to the participants’ diet or physical activity If food intakes or duration of physical activity were **below** the prespecified thresholds, participants would be instructed to increase food consumption to exactly the prespecified portions or increase physical activity duration to 30 minutes per day (when applicable);
3. if changes are required, participants would be instructed to decrease consumption of foods not recommended to the extent of the increase in (2). For example, if a 2-serving increase in vegetables and fruits is required to meet the pre-specified intervention thresholds, participants would be instructed to substitute 2 servings of vegetables and fruits for non-whole grain foods, other low nutritive value foods, juice, sugary drinks and alcohol, and fatty foods rich in saturated fats.

In the emulation, for all hypothetical interventions, the substitution will be implemented by including “total intakes” as a covariate and leaving out foods not recommended from the models. More precisely, a variable reflecting total food intake (in RA/day) and a variable reflecting total beverage intake (in RA/day) will be included in all models.

Hence, total food and beverage intakes will be constant across hypothetical diet interventions. In this approach to account for “total energy”, all model coefficients reflect the action of increasing intakes of recommended foods and a concomitant decrease in *any* of the foods not recommended [32]. On the one hand, this approach is potentially confusing [17,33], since the default interpretation of model coefficients is the action of increasing the intake of each **food included in the model**, while decreasing the intake of foods **not in the model** [32]. On the other hand, the standard model is generally consistent with the implementation of dietary intervention in feeding trials [16,36,37].

The standard model also reduces the number of variables to be considered as intervention variables. Otherwise, 4 additional dietary components would have to be modelled for foods not recommended (i.e., non-whole grain foods, other low nutritive value foods, juice, sugary drinks and alcohol, and fatty foods rich in saturated fats). Finally, the explicit description of the intervention strategies in the target trial protocol clarifies the estimand of interest, as done previously [19,31].

Of note, nutrient-based recommendations in CFG (i.e, saturated fats, free sugars and sodium) are not explicitly modelled to avoid the problems associated with mixed-unit models [17]. In the target trial, we assume that nutrient-based targets would be met by reducing consumption of foods not recommended (i.e., non-whole grain foods, other low nutritive value foods, juice, sugary drinks and alcohol, and fatty foods rich in saturated fats). In that regard, food-level substitution analyses in Canadians support this assumption for saturated fats [38,39].

The “additional protein” intervention consists of increasing the intake of both animal-based and plant-based protein foods by 1.5 and 0.5 RA per day, respectively, as well as milk and plant-based beverages with sufficient protein by 0.5 RA per day. In terms of amount of food, this corresponds to adding 150 grams of cooked unprocessed red meat, fish or poultry or 3 small eggs, 25 grams of nuts and seeds and 125 ml of milk or plant-based beverages with sufficient protein, while proportionally decreasing the intake of foods not recommended. In a previous RCT [40], older women aged 60 to 90 years were able to consume an additional 160 grams of cooked lean red meat without substitution, hereby supporting the feasibility of the protein intervention in the present hypothetical study.

### Assignment

Random allocation (randomization) will be emulated by adjusting for dietary intakes in the year prior to the intervention, as well as adjusting for covariates at the start of the study. Covariates were identified using the causal diagrams depicted in *Figure 1* based on background knowledge of the relationship between the hypothetical lifestyle intervention and outcomes.

**Figure 1:**
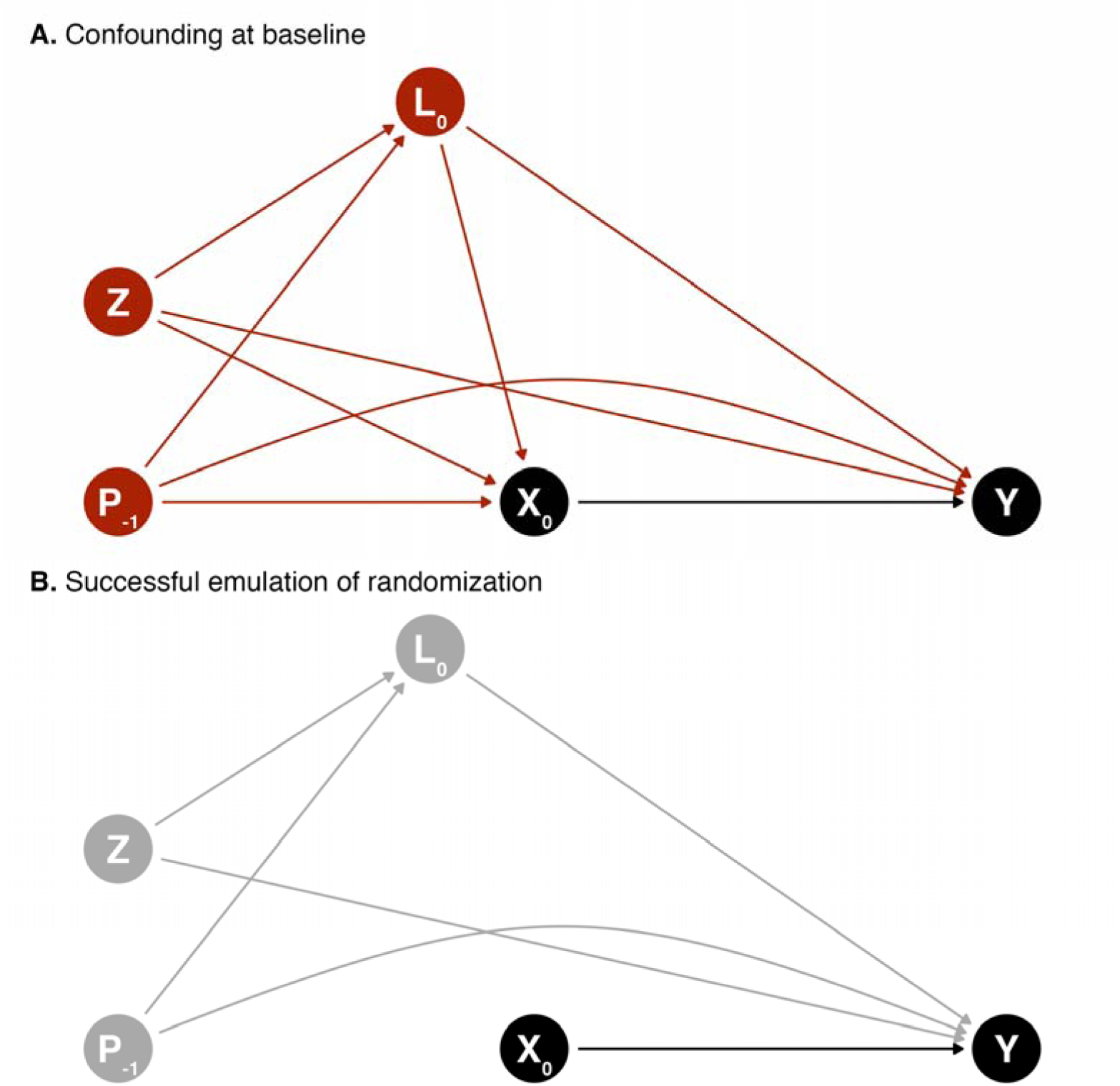
Causal directed acyclic graph (DAG) depicting confounding (A) and successful emulation of randomization using g methods (B) at baseline between the intervention strategy (X) and outcome (Y). Baseline covariates (both time-invariant [Z] and time-varying [L]) as well as prior diet and physical activity habits (P) must be considered to emulate randomization. The successful emulation of randomization of the lifestyle intervention strategy requires no unmeasured confounders and no residual confounding. Time-varying treatment and covariates are not shown in this DAG to focus on randomization emulation.

Dietary components that are the foundation for healthy eating in CFG include intakes of vegetables and fruits, whole grains, protein foods (plant- and animal-based protein foods, milk and plant-based beverages with protein), unsaturated oils and fats [1].

Covariates include age at baseline, biological sex, region, education, living alone, smoking and drinking (alcohol) habits, major chronic diseases (i.e., hypertension, diabetes, cancer, heart disease), number of medications, supplement use (e.g., vitamins and minerals), living alone, and height and weight.

- Z, baseline covariates: age, sex, region, education, history of smoking, height, former cancer.
- P, prior exposure (i.e., exposure of time-varying intervention prior to baseline): dietary habits before baseline
- L, (time-varying) covariates: weight, number of medications, supplement use, living alone, major chronic diseases (hypertension, diabetes, cancer, heart disease), smoking and alcohol habits
- X, (time-varying) treatment: diet and physical activity habits
- Y, end of follow-up outcome: muscle health, general health, cognitive health.

Contrary to dietary habits, data on physical activity habits before baseline were not collected in the NuAge study. In this case, the potential effect of prior physical activity habits will not be accounted for in the models that aim at emulating the sustained physical activity intervention strategy. For the models emulating the sustained diet intervention strategy only, physical activity habits during the study will be used as covariate, hence mitigating the confounding of prior physical activity, at least to some extent.

A successful emulation of randomization requires that there is no unmeasured confounding. However, this is never guaranteed with observational data. Thus, we emphasize our assumptions that 1) the causal graph accurately depicts the relationship under study; and 2) the covariates included are a sufficient set of covariates to address confounding.

### Outcomes

The primary outcomes will be the mean end of follow-up values for muscle strength (i.e., handgrip using vigorimeter, elbow flexor, knee extensor) and physical function (i.e., normal and fast walking, “timed up-and-go”). Outcome values were measured according to standardized protocol in NuAge [25].

For secondary outcomes, mean end of follow-up values for a set of relevant variables will be considered by domains:

- **General health**: waist circumference, blood pressure (systolic, diastolic), blood glucose, estimated glomerular filtration rate;
- **Cognition**: the modified Mini-Mental State Examination (3MS) score.

### Time Zero and Follow-up

In the target trial of a sustained lifestyle intervention, participants would be met at baseline and then regularly to ensure that diet and physical activity habits are consistent with the intervention assigned by the random allocation. The hypothetical diet and physical activity intervention would be assigned and initiated at baseline. In the emulation, annual follow-ups with comprehensive diet, physical activity and covariate data collection are available to emulate the hypothetical intervention. Hence, participants would be followed from study baseline (time 0; i.e., the time at which the intervention strategy would also be assigned and would begin), at each year (time 1 and 2) and until the end of the study (time 3). We also assume that the diet and physical activity habits measured at each follow-up time adequately reflect the habits during the full year.

The end of follow-up outcome measurements will be used to estimate the effect of the sustained lifestyle intervention strategy. Measures of dietary intakes and physical activity throughout the study (i.e., time 0 to time 3) will be used to emulate the sustained lifestyle intervention. Dietary intakes in the year prior to the intervention will be estimated using the frequency questionnaire completed at study baseline (time 0).

Missing covariate data at a given follow-up will be carried forward once, after which participants will be considered as having incomplete follow-up.

### Causal Contrast

The estimand of interest in this study, the target causal effect of a sustained lifestyle intervention strategy, is

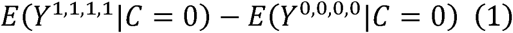

that is, the expected value of a given health outcome Y at the end of follow-up if all participants had increased their adherence to CFG recommendations on healthy food choices and physical activity, when applicable, at all four time points (X_k_ = 1, “always intervene”) vs., instead, if all participants had maintained their habitual diet and physical activity (X_k_ = 0, “never intervene”). The estimand (*Equation 1*) also indicates that all participants completed the intervention (C = 0), i.e., in absence of incomplete follow-up.

The causal contrasts of interest are the observational analogues of “intention-to-treat” and “per protocol” [11]. Given the observational design, participants are not expected to have followed a treatment strategy unknown to them at the time of data collection.

Therefore, the primary analysis will be the per protocol contrast of a sustained lifestyle intervention strategy. In the per protocol analysis, non-adherence to the hypothetical interventions can be accounted for. In the target trial, participants with a condition after baseline that would have prevented or limited participation in a hypothetical lifestyle intervention would be allowed to discontinue the intervention (e.g., lengthy hospitalization, prolonged bed rest, incident cancer). In the emulation, if such conditions occur in a sufficiently large number of participants, these participants will be “excused” from following the hypothetical intervention [23]. In other words, participants that would have been unable to pursue the study due to major events will not be considered as having incomplete follow-up if they attended the annual assessment. Allowing participants to discontinue adhering to the (hypothetical) intervention strategy mitigates confounding by the disease burden [23].

The intention-to-treat analysis will be a secondary analysis of a hypothetical point intervention, e.g., dietary counselling at baseline only. Note that it will not be possible to conduct an intention-to-treat analysis identical to that of a controlled study where the interest is to estimate the effect of being *assigned* to an intervention [11]. However, it is possible to conduct an “observational analogue” of the intention-to-treat analysis. In the observational analogue, the intention-to-treat analysis aims to estimate the impact of a hypothetical intervention which adherence is measured at baseline only.

### Statistical Analysis

Stratification and multivariable regression (i.e., covariate adjustment) are conventional statistical approaches to address confounding in nutritional epidemiology. However, the conventional approaches are not adequate to estimate cumulative treatment effects (e.g., diet over time) in the presence of time-varying confounding (e.g., weight status over time) and treatment (e.g., prior diet) [41,42]. In the present study, non-adherence to the hypothetical interventions and incomplete follow-up will be considered using (general) g methods for the per protocol analysis [11,42]. Among g methods, the parametric g-formula provides the most flexibility for analyses involving hypothetical dietary interventions, as used previously [19,22,43]. Briefly, in the context of an observational study, the parametric g formula, and its implementation into an R package [44], uses parametric models to predict the joint-history of prior diet and physical activity habits (i.e., the hypothetical sustained intervention strategy) and confounding variables. For example, linear regression models are used to predict continuous covariates (e.g., body weight), while logistic regressions are used to predict binary or categorical variables (e.g., indicator variable for dietary supplement use). The per protocol causal contrast of hypothetical intervention presented in Table 2 is then emulated based on Monte Carlo simulated data generated using the g formula algorithm [44]. The parametric g formula correctly accounts for time-varying confounding in the presence of feedback between the intervention and the confounding variables, since confounding is addressed using standardization [41,42]. Furthermore, standardization allows to estimate an average causal effect (i.e., marginal effect) consistent with the estimand of interest (Equation 1) rather than a conditional effect. In summary, a “threshold interventions” that depend on the reported dietary intakes or amount of physical activity [43,45] and the parametric g-formula [44,46] will be used to emulate the intervention of “consuming at least x servings of food” and “doing at least x minutes of light to vigorous physical activity”.

*Figure 2* presents the causal DAG of the hypothesized relationship between a sustained lifestyle intervention strategy (X_0_, X_1_) and an end of follow-up outcome Y, for one follow-up after baseline (year 1). The model is limited to year 1 for clarity but the hypothesized causal structure extends to additional follow-ups. The exposure of interest X_k_ is the joint and cumulative effect of a sustained diet and physical activity intervention strategy measured at baseline and at follow-ups.

**Figure 2:**
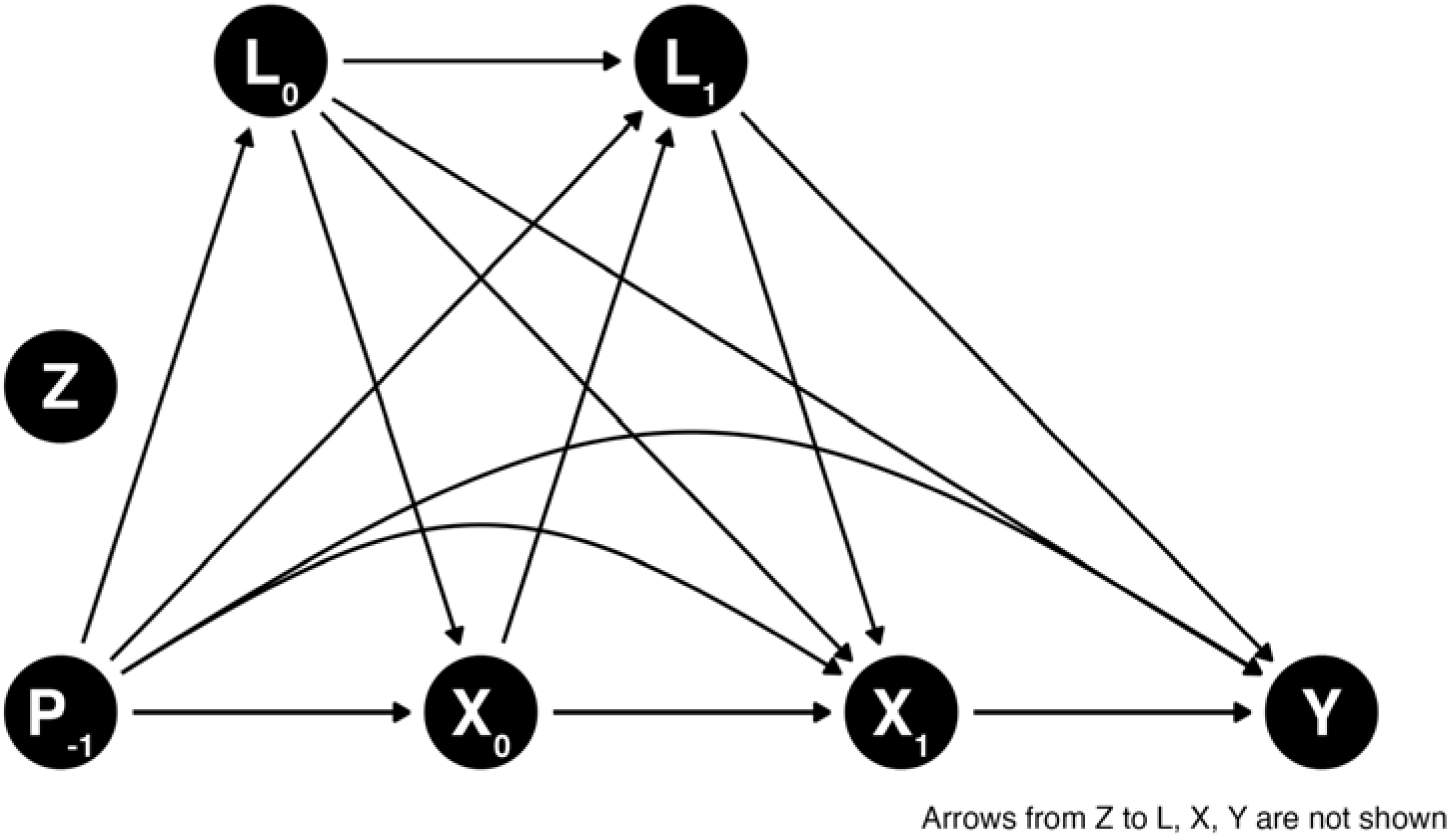
Directed acyclic graph (DAG) depicting the hypothesized relationship among prior dietary exposure and time-varying interventions (diet, physical activity) and covariates. Arrows from the Z node are not shown for clarity, but would point towards all baseline and time-varying nodes, as well as outcome. Only one follow-up is shown for visualization purpose, but the hypothesized causal structure extends to additional follow-ups.

In the context of this target trial emulation (Figure 2), P includes dietary habits prior to the baseline assessment. P can have an effect on baseline dietary habits (e.g., prior healthy habits increase likelihood of baseline healthy habits) and dietary habits throughout the target trial emulation (e.g., prior healthy habits increase likelihood of adhering to healthy habits). P also influences baseline and time-varying confounding.

Finally, given the long-term effect of chronic exposure, P potentially also affect Y directly.

The intention-to-treat analysis is similar to the per protocol analysis. However, only baseline diet and physical activity habits and covariates are considered as well as pre-baseline diet and physical activity. In both the per protocol and intention-to-treat analyses, loss to follow-up (e.g., non-response or missing follow-up, health outcomes not measured) will be accounted for using g-methods such as inverse probability weighting.

#### Dietary Assessment

In the NuAge study, diet during the year before baseline was assessed using one semi-quantitative food-frequency questionnaire [47]. Dietary intakes at baseline (time 0) and each annual follow-ups (time 1, 2 and 3) were assessed using 3 repeated face-to-face interviewer-administered 24-hour dietary recall.

Dietary intakes measured using 24-h dietary recall are more accurate (i.e., less systematic error or bias) than food-frequency questionnaire [48–50] but are particularly affected by random measurement error (i.e., within-individual random error) [50]. When several variables measured with errors are considered simultaneously in a regression model, the regression coefficients may be biased in any direction [51]. To account for random measurement error, the National Cancer Institute Markov Chain Monte Carlo (MCMC) multivariate method could be applied [52]. However, the combination of the parametric g-formula and multivariate measurement error correction using the NCI MCMC method is not feasible. The NCI MCMC method estimates time-invariant measurement error-corrected intakes, while the g formula algorithm is designed for time-varying exposures.

Recognizing the importance of accounting for measurement error, the correction for measurement error will be reserved for the secondary intention-to-treat analysis. For the intention-to-treat contrast, the time-varying values of exposure and the time-varying values of confounding are not considered. Thus, the NCI MCMC method will be used to obtain measurement error-corrected estimates of the relationship between dietary intakes measured at baseline and outcome at the end of follow-up. Of note, the three 24-h dietary recalls collected at each time point contribute to reducing random errors, at least to some extent, even in absence of measurement error correction.

#### Sensitivity analysis to assess the impact of measurement error

For the secondary intention-to-treat analysis, results based on the measurement error-corrected and uncorrected dietary intakes will be compared. The difference between the estimated relationships will allow to extrapolate the impact of not accounting for random errors in the primary *per protocol* analysis.

#### Physical activity assessment

Physical activity throughout the study was assessed using the Physical Activity Scale for the Elderly (PASE) questionnaire [53,54]. Rather than the total PASE score, specific questions estimating the total time of physical activities will be used to be consistent with the intervention strategy.

#### Covariates and subgroups

To the extent permitted by the number of observations for each outcome, continuous covariates will be modelled using restricted cubic splines with 3 to 5 knots (at percentiles 10-50-90, 5-35-65-95 or 5-27.5-50-77.5-95) [55]. Categorical covariates will be modelled to ensure a sufficient sample size in each level.

The effect of dietary changes will be estimated for the full sample. The sample will also be stratified by (biological) sex to reflect both potential biological differences and, to some extent, gender differences (although not reported).

#### Variance estimation

Variance will be estimated using a minimum of 200 bootstrap sample replicates to consider uncertainty at each step of the estimation [56].

#### Software and code

The main statistical analyses will be conducted using R (version 4.3.1 or greater) and the gfoRmula package [44,57]. The manuscript results will be generated using Quarto markdown. Codes for main analyses and generation of manuscript results will be shared in a publicly available code repository.

## Ethical Considerations

### Human subject ethics review approvals or exemptions

The original NuAge study protocol was approved by the Research Ethics Boards of the *Institut universitaire de gériatrie de Montréal* and the *Institut universitaire de gériatrie de Sherbrooke* (Quebec, Canada). The NuAge Database and Biobank [58] has received approval by the Research Ethics Board of the *Centre intégré universitaire de santé et de services sociaux de l’Estrie—Centre hospitalier universitaire de Sherbrooke*. Secondary analyses of data from the NuAge Database and Biobank for the study described in this protocol are approved by the McGill University Research Ethics Board Office (#22-11-041).

### Informed Consent

All participants of the NuAge study provided informed consent. From the initial cohort of 1,793 participants, 1,753 (98%) agreed to the integration of their data and biological samples into the NuAge Database and Biobank for future studies.

### Privacy and confidentiality

Secondary analyses based on the NuAge Database and Biobank use de-identified data, which do not allow participants to be identified by the investigators.

### Compensation details

Participants from the NuAge study voluntarily consented to participate and were not provided with monetary compensation.

## Results

Data collection for the NuAge study was completed in June 2008. For the present work, the main analysis based on the final curated data started in May 2024. The manuscript will be written according to the Strengthening the Reporting of Observational Studies in Epidemiology (STROBE) statement. We anticipate the submission of the manuscript to peer-reviewed academic journal by December 2024.

## Discussion

In this study protocol, we have described a target trial to assess the effect of adhering to CFG’s recommendations on healthy food choices. The emulation will be performed using data from the NuAge Database and Biobank [58]. Benefiting from the flexibility of observational data, we also aim to compare adherence to multiple reformulations of CFG’s recommendations, including the effect of increasing the intake of protein-rich foods and the amount of aerobic physical activity on selected health outcomes. We have also described the rationale for using simulated diets to emulate the adherence to CFG’s recommendations, the process of selecting covariates - to attempt - to emulate randomization with causal diagrams, and the challenges of addressing random measurement error.

We emphasize that the purpose of emulating a target trial using observational data is to improve the quality of observational analysis [11,14]. In other words, the target trial framework aims to support the coherence between the (causal) research question and the observational data analysis [26]. However, estimating causal effects with non-experimental observational data depends on strong assumptions. The key assumptions are that there are no unmeasured confounder, no measurement error and no model misspecification (e.g., functional form of covariates, model outcome distribution) [19].

We first recognize that the absence of residual or unmeasured confounding cannot be guaranteed. The extent to which this assumption is sufficiently satisfied depends on the appraisal of covariates considered. In that regard, we have used graphical tools, DAG, to explicitly describe our analytical assumptions and to identify confounding variables [59–61]. Second, the absence of measurement error assumption will not be satisfied considering the use of dietary intake data measured with 24-h dietary recalls. On one hand, 24-h dietary recalls have the least systematic error (bias) compared with other common instruments such as food-frequency questionnaire [48,49]. On the other hand, 24-h dietary recalls are largely affected by within-individual random errors [50], which can cause bias in any direction in multivariable models [51], as in the present study.

This issue is mitigated, at least to some extent, by using average data from three repeated 24-h dietary recalls at each follow-up. Sensitivity analyses comparing estimates based on measurement-error corrected and uncorrected baseline dietary intakes will be used to assess the impact of random measurement errors. Third, the absence of model misspecification will be assessed by examining differences between the observed value of time-varying covariates and the predicted value of time-varying covariates as modeled with the g-formula.

## Limitations

Strengths of this study and protocol include the explicit emulation of a hypothetical trial, the thorough description of the emulation of the sustained dietary intervention, and the use of background knowledge and DAG to derive a sufficient set of confounders.

Limitations must be addressed. First, the sample size of the NuAge study is relatively limited (n=1753), although comprehensive nutrition and covariate data was collected. Second, the target food intakes based on diet simulations from Health Canada *exceed* the 99^th^ percentile of the usual intake distribution of Canadians 65 years or older from Canada in 2015 [3]. The *a posteriori* revision of the dietary intervention targets will be needed if observed dietary intakes in NuAge are too far from targets (Table 2), as was done in a previous nutrition target trial emulation [22]. Third, the presence of random measurement error associated with 24-h dietary recalls may bias estimates. Finally, the target trial emulation cannot replace an actual RCT. Evidence from RCT will be required to confirm the value of either CFG recommendations or the enhanced CFG recommendations.

## Conclusion

In conclusion, the target trial framework is relevant to estimate the causal effect of adhering to CFG’s recommendations using non-experimental data when a RCT is impractical [11,19]. Coupled with key assumptions, including the absence of unmeasured confounding, the absence of measurement error and no model misspecification, we believe the emulation will provide timely evidence regarding the effect of adhering to CFG’s recommendations in older adults and inform on the added value of a reformulation.

## Acknowledgements

Authors’ contributions

Conceptualization: all authors; Methodology: DB and SC; Data curation, formal analysis, visualization and writing – original draft: DB; Supervision: SC; Writing – review & editing: all authors.

## Conflicts of Interest

DB was a casual employee of Health Canada (2019-2020) and held a doctoral training award from the Fonds de recherche du Québec – Santé (2019-2021). DB has no conflicts of interest. SC receives research funding from the CIHR, *Fonds de recherche du Québec*, Canadian Foundation for Dietetics Research, Canadian Foundation for Innovation and Canadian Cancer Society. None of these agencies has funded nor was involved in this work.

NP is the NuAge Database Administrator; NP and SC serve as NuAge Steering Committee Members.

The NuAge Study was supported by a research grant from the Canadian Institutes of Health Research (CIHR; MOP-62842). The NuAge Database and Biobank are supported by the Fonds de recherche du Québec (FRQ; 2020-VICO-279753), the Quebec Network for Research on Aging, a thematic network funded by the Fonds de Recherche du Québec - Santé (FRQS) and by the Merck-Frosst Chair funded by La Fondation de l’Université de Sherbrooke.

## Data availability and code

The data and code used for this manuscript will be made available at https://github.com/didierbrassard/NuAge_protocol

**Multimedia Appendix 1**: Supplemental Table 1. HEFI-2019 dietary constituents and score among simulated diets by Health Canada, by age and sex group

## Abbreviations

CFG: Canada’s Food Guide
DAG: directed acyclic graph
HEFI: Healthy Eating Food Index 2019
MCMC: Markov Chain Monte Carlo
NCI: National Cancer Institute
RA: Reference amount
RCT: randomized controlled trial

