## Supplemental Table 1 for "Estimating The Effect Of Adhering To Canada’s Food Guide 2019 Recommendations On Health Outcomes In Older Adults: A Target Trial Emulation Protocol"

Supplemental Material

Didier Brassard

Nancy Presse

Stéphanie Chevalier

### Supplemental Methods

#### Simulated diets from Health Canada

Table 1: Supplemental Table 1. HEFI-2019 dietary constituents and score among simulated diets by Health Canada, by age and sex group*^1^*

|  | Foods, Reference Amounts (RA) | | | | | | Total nutrients | | | | |  |
| --- | --- | --- | --- | --- | --- | --- | --- | --- | --- | --- | --- | --- |
| Age/sex group | Vegetables & fruits | Whole grains | Protein foods, plant-based | Protein foods, animal-based | Milk & Plant-based bev. with protein | Unsaturated oils & fats | Energy, kcal | Free sugars, g | SFA, g | Unsaturated fats, g | Sodium, mg | HEFI-2019 score (/80), points*^2^* |
| All, 51y+ | 9.0 | 3.9 | 2.0 | 1.9 | 1.0 | 1.0 | 1,865 | 6.4 | 10.2 | 41.6 | 1,066 | 78.5 |
| Males, 51-70 y | 9.9 | 5.0 | 2.4 | 2.3 | 1.0 | 1.0 | 2,259 | 7.1 | 11.7 | 45.2 | 1,174 | 78.3 |
| Females, 51-70 y | 8.6 | 3.5 | 2.0 | 1.9 | 1.0 | 1.0 | 1,737 | 5.5 | 9.7 | 41.3 | 993 | 78.3 |
| Males 71y+ | 9.9 | 4.2 | 2.0 | 1.8 | 1.0 | 1.0 | 1,959 | 7.0 | 10.6 | 43.0 | 1,142 | 78.6 |
| Females 71y+ | 7.7 | 3.0 | 1.7 | 1.6 | 1.0 | 1.0 | 1,507 | 5.9 | 8.7 | 36.8 | 954 | 78.0 |
| *^1^*Data adapted from Health Canada. Simulated composite diets. https://open.canada.ca/data/en/dataset/0490749d-b0b0-410a-9577-a903c6cec2be: Open Government Portal, 2022. CFG-2019, Canada's Food Guide; DRI, Dietary Reference Intake; HEFI-2019, Healthy Eating Food Index 2019; RA, reference amounts; SFA, saturated fats. | | | | | | | | | | | | |
| *^2^*The HEFI-2019 total score has a maximum of 80 points. Foods not recommended (i.e., non-whole grain foods, other low nutritive value foods, juice, sugary drinks and alcohol, and fatty foods rich in saturated fats) were all assigned 0 consumption. | | | | | | | | | | | | |
